# Exploring the Potential of Large Language Models for Automated Safety Plan Scoring in Outpatient Mental Health Settings

**DOI:** 10.1101/2025.03.26.25324610

**Authors:** Hayoung K. Donnelly, Gregory K. Brown, Kelly L. Green, Ugurcan Vurgun, Sy Hwang, Emily Schriver, Michael Steinberg, Megan Reilly, Haitisha Mehta, Christa Labouliere, Maria Oquendo, David Mandell, Danielle L. Mowery

## Abstract

The Safety Planning Intervention (SPI) produces a plan to help manage patients’ suicide risk. High-quality safety plans – that is, those with greater fidelity to the original program model – are more effective in reducing suicide risk. We developed the Safety Planning Intervention Fidelity Rater (SPIFR), an automated tool that assesses the quality of SPI using three large language models (LLMs)—GPT-4, LLaMA 3, and o3-mini. Using 266 deidentified SPI from outpatient mental health settings in New York, LLMs analyzed four key steps: warning signs, internal coping strategies, making environments safe, and reasons for living. We compared the predictive performance of the three LLMs, optimizing scoring systems, prompts, and parameters. Results showed that LLaMA 3 and o3-mini outperformed GPT-4, with different step-specific scoring systems recommended based on weighted F1-scores. These findings highlight LLMs’ potential to provide clinicians with timely and accurate feedback on SPI practices, enhancing this evidence-based suicide prevention strategy.

## Introduction

The Safety Planning Intervention (SPI) is a widely used, evidence-based intervention to prevent suicide. It is designed to help patients develop personalized strategies for managing suicide crises. SPI consistently demonstrates effectiveness in reducing suicide risk across various settings, including emergency departments^1^, outpatient care^2^, and community contexts such as the Department of Veteran Affairs^3^. Unfortunately, most evidence suggests that clinicians do not implement SPI as it was designed. Prospective studies have found that lower-quality safety plans are associated with increased risk of suicidal behavior^4^.

Providing real-time feedback to clinicians could greatly enhance their use of SPI^5-7^. The current state of the science would require direct observation by expert clinicians to provide this feedback, however. Even asynchronous feedback on SPI implementation is challenging. Manually evaluating and providing feedback on safety plan quality requires significant time and specialized training^8^. Clinical decision support systems could provide clinicians with timely and specific feedback for improving fidelity to SPI, and ultimately, reducing patient suicide risk. In this pilot study, we developed and pilot tested an automated Safety Planning Intervention Fidelity Rater (SPIFR), a tool for scoring safety plans and providing real-time feedback to clinicians. We developed the SPIFR by comparing three large language models (LLMs) based on their weighted F1 scores and optimizing their prompts and parameters to maximize performance. We hypothesized that o3-mini would outperform GPT-4 and LLaMA 3 in effectively scoring safety planning templates when compared to scores assigned by trained clinical coders.

## Methods

This study was approved by the University of Pennsylvania IRB with the data use agreement and transfer from Columbia University.

### Data

We analyzed 266 deidentified safety plan forms collected as part of a Zero Suicide implementation project^9^. Safety plan forms were collected from patients who received SPI across 61 outpatient mental health clinics in New York State. Narrative text on the safety plan is collaboratively developed by the clinician and the patient. Coders assessed safety plan narrative quality using the Safety Planning Intervention Scoring Algorithm (SPISA)^10^. The SPISA consists of 20 items measuring the quality of narrative responses across seven steps of the safety plan form: w*arning signs, internal coping strategies, social contacts and social settings, social contacts for a crisis, professionals for a crisis, making the environment safe*, and *reasons for living*. The original SPISA includes two measures that assess the quality and completeness of safety plan responses. **Quality** of response evaluates how specific and personalized a patient’s response is as well as its relevance to each step’s question. **Completeness** of response measures whether patients answered all steps and questions. We focused on developing an automated scoring tool for the quality of responses, particularly for the following four steps of the Safety Plan: w*arning signs, internal coping strategies, making the environment safe*, and *reasons for living*. The remaining three steps were excluded due to a high number of missing entries following the deidentification process, as they often contained personal information (e.g., names, phone numbers) that were redacted in compliance with the Health Insurance Portability and Accountability Act (HIPAA) of 1996 using the Safe Harbor method.

Below are the definitions of the four target steps along with fictitious, but realistic examples:

- *Warning signs:* specific thoughts, feelings, physiological states, or behaviors that are associated with the development of a suicidal crisis (e.g., “not sleeping well”, “my heart starts pounding faster”)
- *Internal coping strategies:* activities or behavior people can engage in by themselves that distract from suicidal thinking and help them feel better (e.g., “write in my journal”, “listening to the Beatles”)
- *Making the environment safe:* an action plan that the client should take to reduce access to lethal means (e.g., “get rid of unused medication”, “lock up my firearm and give the key to my brother”)
- *Reasons for living*: things that matter most to clients and give them a sense of purpose and motivation to continue living (e.g., “my family”, “I want to go to college, become a nurse, and help people”)

The possible maximum number of responses differed for each step: 3 for *warning signs*, 3 for *internal coping strategies*, 2 for *making the environment safe*, and 1 for *reasons for living*. Among 2,210 individual responses across four steps from the 266 safety plan forms, we observed 772 responses for *warning signs*, 770 responses for *internal coping strategies*, 405 responses for *making the environment safe*, and 263 responses for *reasons for living*.

In **Figure 1**, we present a workflow diagram illustrating an example of the SPI, the SPIFR development and output.

**Figure 1.**
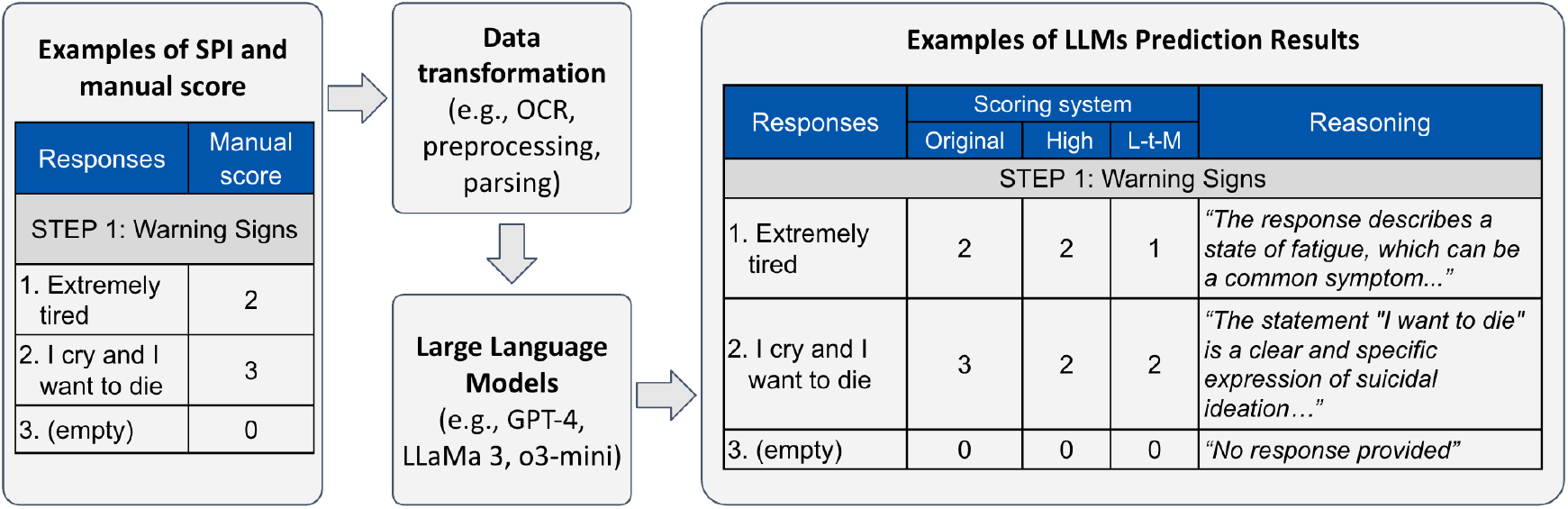
Workflow of the SPIFR development

### OCR (Optical Character Recognition)

The original data files are PDF documents containing typed safety planning text without handwritten content. We used Tesseract^11^, an open-source text recognition engine, to extract text for large language model development. After converting the PDF files to TIFF format using the python imaging library, we applied Tesseract version 5 with default configurations for English text extraction, generating plaintext output.

### Parsing

The original dataset consists of various text elements beyond patients’ narrative texts including step titles, operational definitions of concepts (e.g., “Internal coping strategies - things I can do to take my mind off my problems without contacting another person (distracting & calming activities)”), and other information (e.g., page numbers). First, to focus solely on the responses for each step, these elements were removed from the data. Second, for documents with explicit step numbering (e.g., “Step 1. Warning signs” and “Step 2: Internal coping strategies”), the text between these markers were extracted to capture content related to each specific step. Third, there were cases where these patterns were not found. Instead, the documents presented items like “Warning signs: 1. (response 1) 2. (response 2) … Internal Coping Strategies: 1. (response 1) 2. (response 2) …” In such cases, the text between the first and second occurrence of “1.” (or “1:”) was extracted as the response for step 1 warning signs. Similarly, the text between the second and third occurrence of “1.” (or “1:”) was extracted as the response for step 2 internal coping strategies. Fourth, for the final step (e.g., “Step 7: Reasons for living”), which lacks a clear ending marker (such as a “Step 8”), other textual indicators that follow the last step (e.g., “The Stanley-Brown Safety Plan is copyrighted by…”, “Patient signature:”) were used to determine the endpoint of the responses. Finally, after extracting the responses for each step, they were further processed, because each individual response requires separate scoring. If the responses were numbered (e.g., “1. 2. 3.”), this numbering was utilized to split and identify each response individually. In instances where the responses were not numbered, the text was divided by newline characters, and each line was treated as a distinct response.

### Prompt Development and Scoring Algorithm

We employed a zero-shot prompting strategy to rate each response and compared model performance of three scoring systems. The prompt was developed based on the SPISA scoring algorithm manual and iteratively refined to enhance the model’s performance throughout the process. We tested three scoring systems: the **original** scoring system based on SPISA, a **high** precision system, and a **low-to-moderate** precision system. The 4-score **original** system consists of four levels: (0) no response, (1) general, (2) somewhat specific, and (3) highly specific. Examples of responses with varying levels of specificity and corresponding scores are provided in **Figure 2a**. The 3-score **high** precision system has (0) no response, (1) general and (2) somewhat and highly specific. The 3-score **low-to-moderate** precision system has (0) no response, (1) general and somewhat specific and (2) highly specific responses. To improve the explainability of the rater^12^, we also incorporated reasoning generation for each score, ensuring that clinicians receive interpretable feedback.

**Figure 2a.**
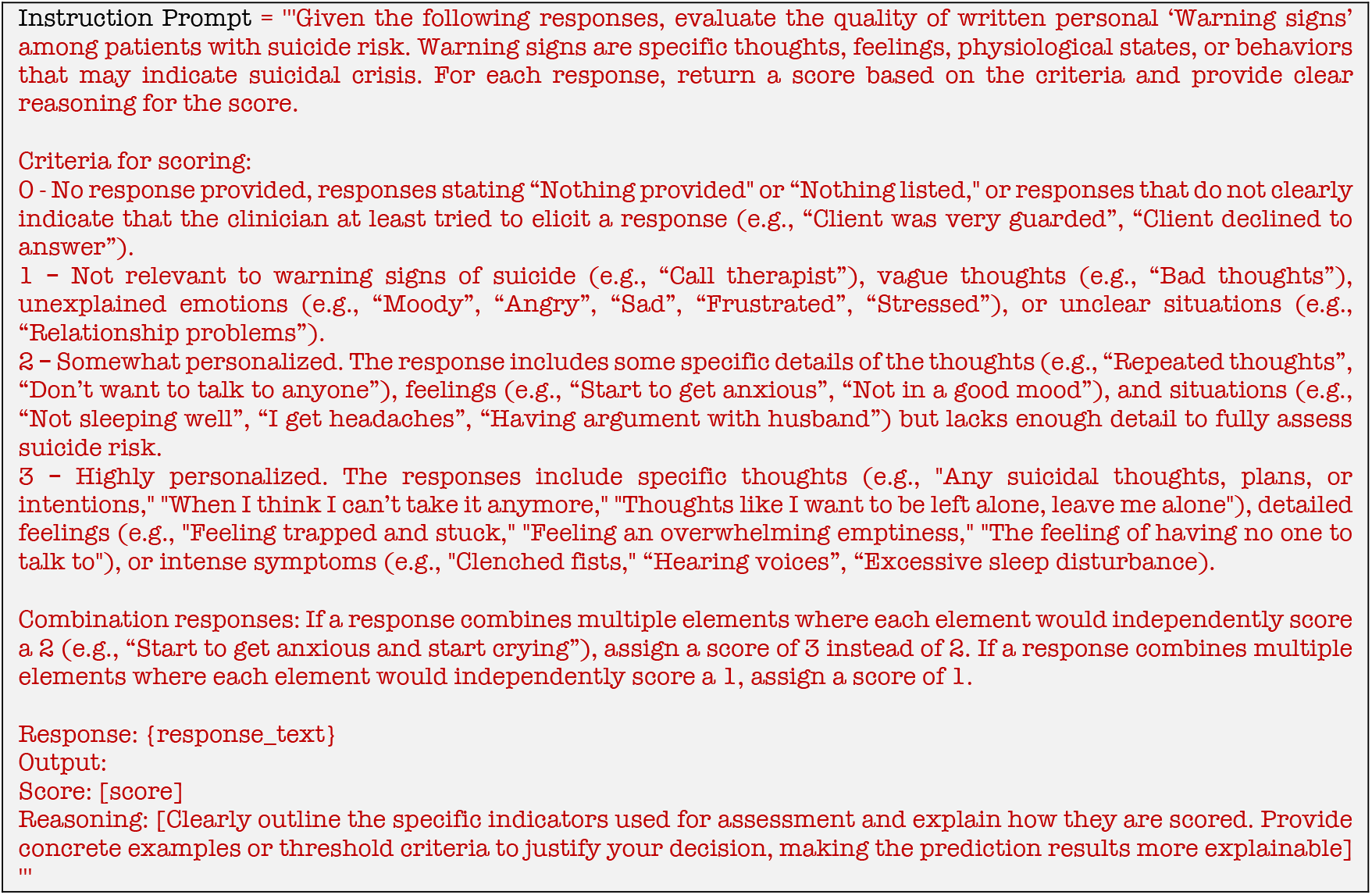
Prompt example for step 1: warning signs

**Figure 2b.**
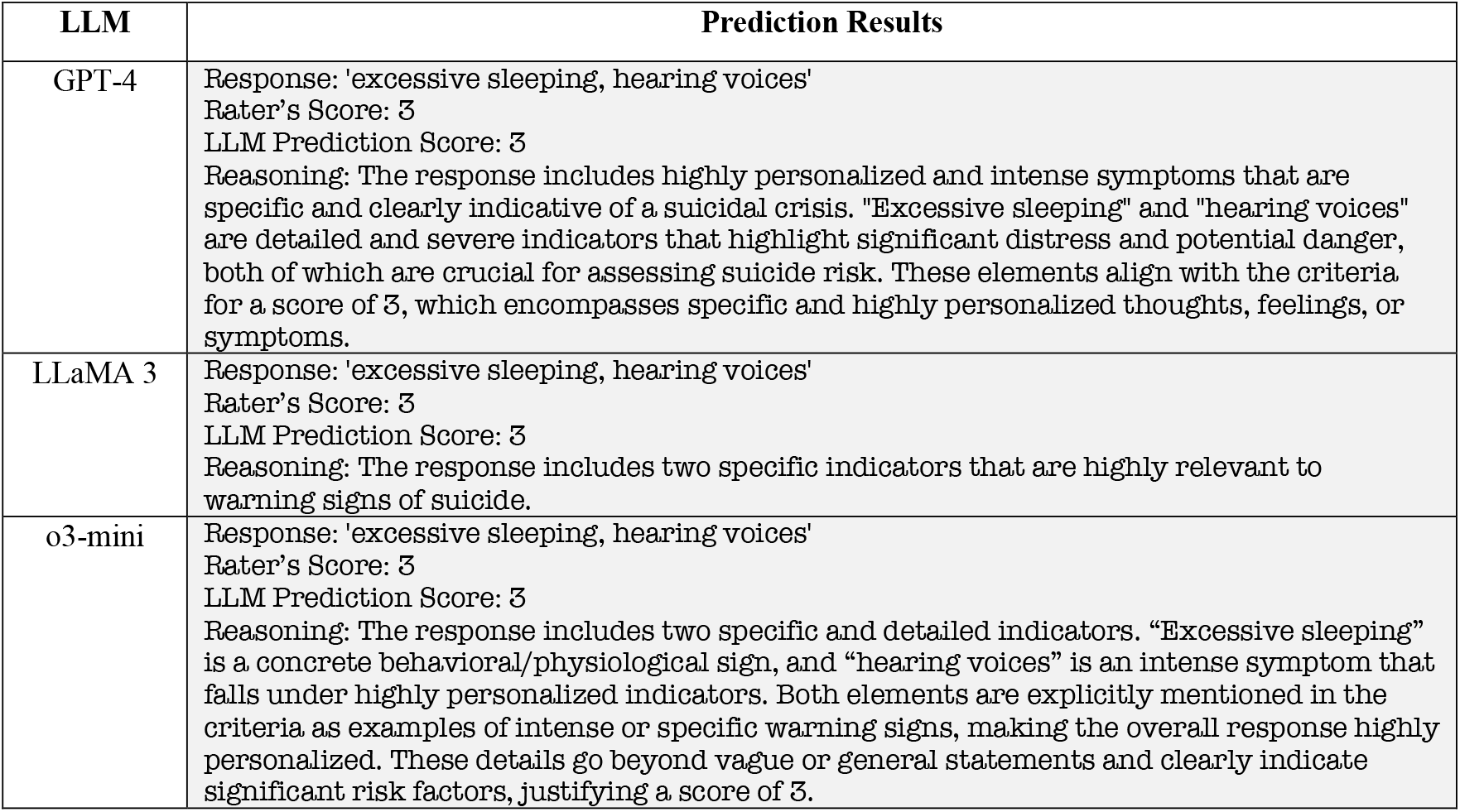
Examples of prediction results with reasoning for Step 1: warning signs for each LLM.

### Large Language Models

Using Microsoft Azure Databricks, we assessed the performance of the SPIFR tool using the SPISA-coded data and three LLMs: GPT-4, LLaMA 3, and o3-mini. We originally selected only GPT-4 because it tends to perform clinical coding tasks well, but we wanted to understand whether an open-weight model like LLaMA 3 was comparable in performance and o3-mini’s reasoning ability outperformed this baseline model. For GPT-4 and LLaMA 3, we compared models using three temperature settings [0.1, 0.5, 0.9]. For o3-mini, we evaluated performance across three reasoning effort levels [low, medium, high]. Four steps of safety plan were evaluated by comparing the weighted F1-score (a measure of how accurately a model makes predictions by balancing the impact of different types of prediction errors and ensuring that categories with more data have a greater influence on the final score). We selected F1-score because it is a recommended metric especially for multi-class prediction with imbalanced data^13^, providing more reliable measure of model performance by balancing both precision and recall.

In **Figure 2a**, we provide an example prompt for step 1 *warning signs* using the original 4-value SPISA scoring system and **Figure 2b** for the associated predictions and reasoning for each of the three LLMs.

## Results

### Data Characteristics

The number of responses coded by professionals, with scores of 0, 1, 2, or 3, displayed an asymmetrical distribution (**Figure 3**). For *warning signs* response (n = 772), the highest proportion of responses (38.99%, n = 301) received a score of 2, followed by 38.6% (n = 298) scored 3, 17.88% (n = 138) scored 1, and 4.53% (n = 35) scored 0. For *internal coping strategies* (n = 770), 44.94% (n = 346) of responses scored 1, followed by 29.87% (n = 230) scoring 2, 19.61% (n = 151) scoring 3, and 5.58% (n = 43) scoring 0. Among responses of *making the environment safe* (n = 405), 36.54% (n = 148) scored 1, followed by 28.64% (n = 116) scored 0, 24.69% (n = 100) scored 2, and 10.12% (n = 41) scored 3. Lastly, for *reasons for living* (n = 263), most respondents (69.96%, n = 184) scored 2, with 15.21% (n = 40) scoring 3, 8.37% (n = 22) scoring 0, and 6.46% (n = 17) scoring 1.

**Figure 3.**
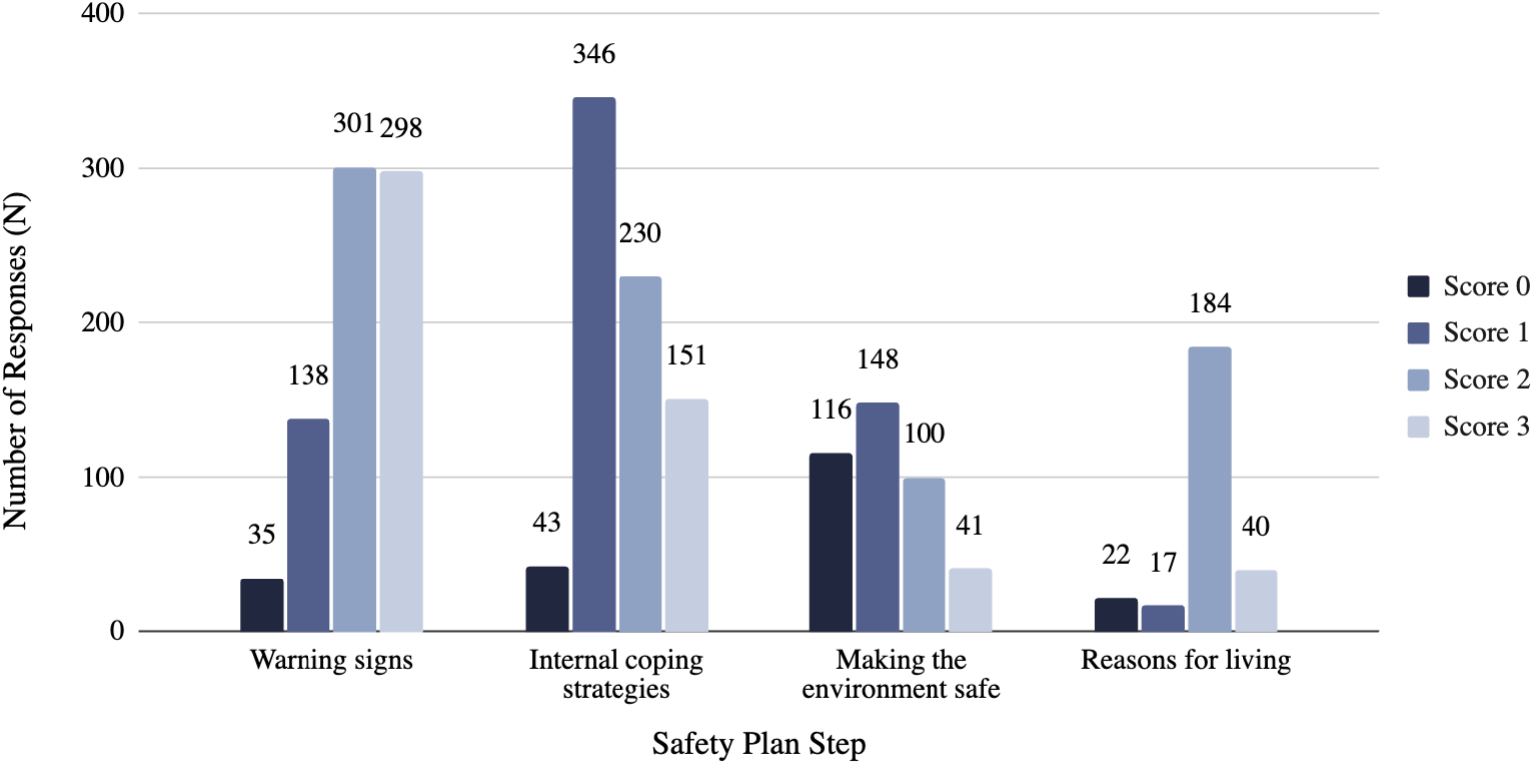
Distribution of responses by quality scores coded by trained raters

The prediction model for the quality scores of safety plan responses varied according to different scoring systems and LLMs (**Table 1**). We assessed four key questions: 1) how well do LLMs score the original scoring system? 2) do LLMs have improved performance with high and low-to-moderate precision scoring systems? 3) does one LLM outperform the others more consistently across steps? and 4) which LLM is most consistent in its ratings?

**Table 1.** Weighted F1-score (%) across large language models. Mean refers to the average weighted F1-score across different hyperparameters (e.g., temperature for GPT-4 and LLaMA 3, reasoning effort for o3-mini); An asterisk (*) indicates the best-performing model for each step. L-t-M refers to the **low-to-moderate** precision system scale.

| SPI step | Scoring system | GPT-4 |  |  |  | LLaMA 3 |  |  |  | o3-mini |  |  |  |
| --- | --- | --- | --- | --- | --- | --- | --- | --- | --- | --- | --- | --- | --- |
|  |  | Mean | Min | Max | Range | Mean | Min | Max | Range | Mean | Min | Max | Range |
| Step 1<br>warning signs | Original | 48.93 | 48.57 | 49.23 | 0.66 | 51.78 | 51.47 | 52.22 | 0.75 | 38.12 | 36.66 | 40.30 | 3.64 |
|  | High | 73.51 | 72.11 | 74.71 | 2.60 | 78.68 | 78.45 | **79.12\*** | 0.67 | 59.53 | 59.03 | 60.46 | 1.43 |
|  | L-t-M | 63.76 | 63.29 | 64.39 | 1.10 | 65.39 | 65.11 | 65.94 | 0.83 | 63.04 | 61.84 | 64.02 | 2.18 |
| Step 2<br>internal coping strategies | Original | 53.27 | 52.70 | 54.08 | 1.38 | 60.19 | 59.59 | 60.65 | 1.06 | 58.86 | 56.56 | 60.73 | 4.17 |
|  | High | 64.22 | 63.52 | 64.85 | 1.33 | 72.77 | 72.11 | 73.18 | 1.07 | 70.50 | 68.67 | 72.57 | 3.90 |
|  | L-t-M | 76.92 | 76.08 | 77.94 | 1.86 | 79.33 | 79.13 | 79.53 | 0.40 | 80.15 | 78.30 | **81.13\*** | 2.83 |
| Step 6<br>making environment safe | Original | 56.84 | 56.32 | 57.24 | 0.92 | 58.52 | 58.35 | 58.68 | 0.33 | 59.73 | 58.52 | 60.52 | 2.00 |
|  | High | 64.33 | 63.92 | 64.72 | 0.80 | 67.38 | 67.16 | 67.61 | 0.45 | 67.90 | 66.86 | 68.55 | 1.69 |
|  | L-t-M | 75.01 | 73.61 | 75.73 | 2.12 | 74.40 | 73.74 | 74.82 | 1.08 | 74.38 | 72.96 | **76.56\*** | 3.60 |
| Step 7<br>reasons for living | Original | 75.79 | 73.65 | 77.22 | 3.57 | 78.86 | 78.38 | 79.52 | 1.14 | 74.47 | 69.26 | 78.81 | 9.55 |
|  | High | 88.23 | 85.67 | 89.97 | 4.30 | 90.69 | 90.38 | 91.02 | 0.64 | 91.72 | 91.42 | **91.99\*** | 0.57 |
|  | L-t-M | 81.61 | 79.52 | 82.71 | 3.19 | 84.21 | 84.15 | 84.34 | 0.19 | 80.56 | 75.39 | 84.44 | 9.05 |

*How well do LLMs score the original scoring system?* The mean performance for **original** scoring systems ranged by steps: *warning signs:* 38.12-51.78, *internal coping strategies:* 53.27-60.19, *making environment safe:* 56.84-59.73, and *reasons for living*: 74.47-78.86.

*Do LLMs have improved performance with high and low-to-moderate precision scoring systems?* We observed increases in predictive power with augmented precision scoring systems. The most elevated mean performance by scoring systems ranged by steps: *warning signs* (**high**): 59.53-78.68, *internal coping strategies* (**low-to-moderate**): 76.92-80.15, *making environment safe* (**low-to-moderate**): 74.38-75.01, and *reasons for living* (**high**): 88.23-91.72.

*Does one LLM outperform the others more consistently across steps?* LLaMA 3 produced the highest predictive performance for *warning signs* (**high**): 79.12; O3-mini produced the highest predictive performance for *internal coping strategies* (**low-to-moderate**): 81.13, *making environment safe* (**low-to-moderate**): 76.56, and *reasons for living* (**high**): 91.99

*Which LLM is most consistent in its ratings?* LLaMA 3 (0.72) produced the most consistent performance, with the smallest mean range difference across different parameter values, compared to GPT-4 (1.99) and o3-mini (3.72).

### Best Rating Model for Each Step

The highest-performing model for each step was as follows (**Figure 4**):

**Figure 4.**
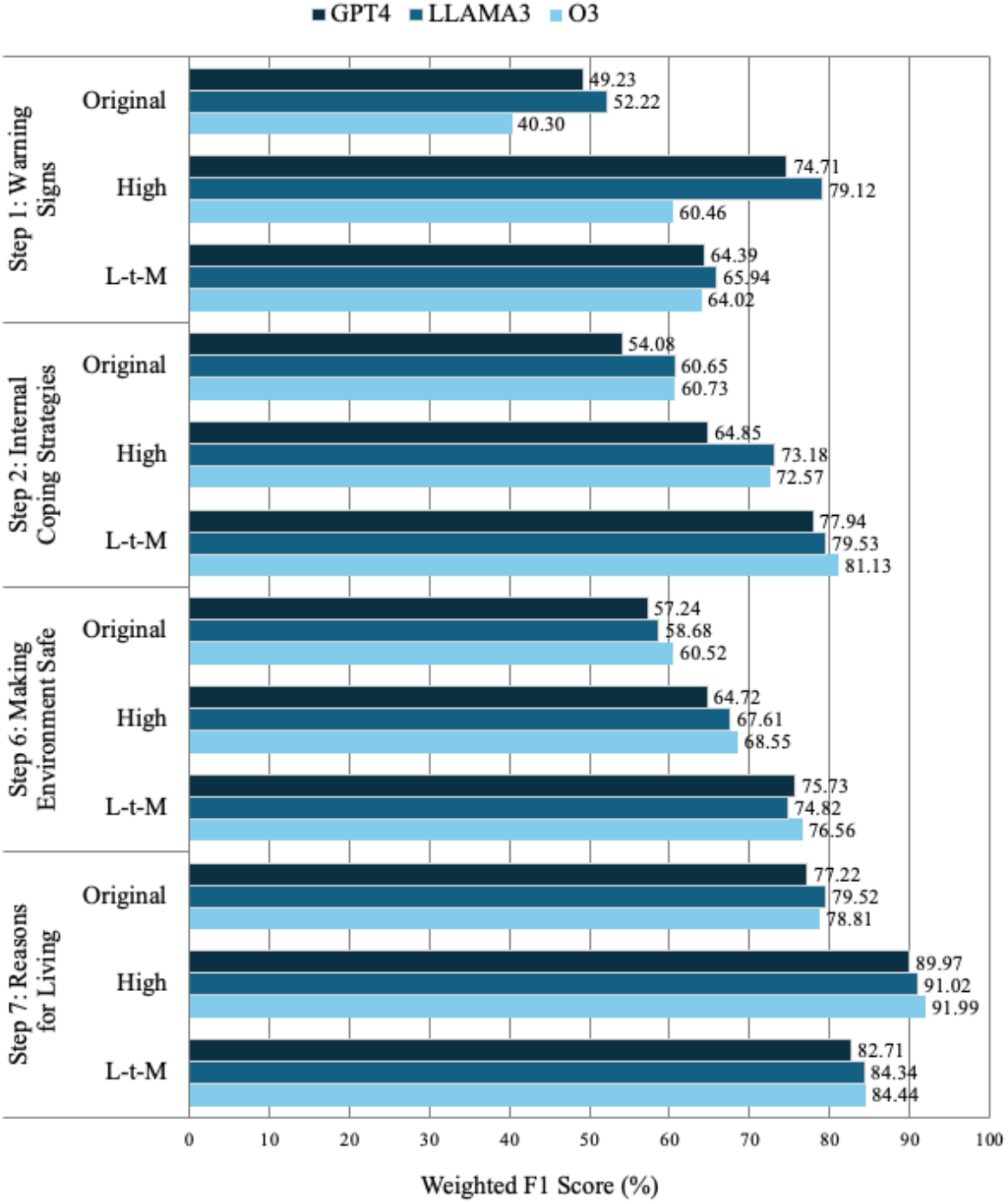
Best model performance across steps, scoring systems, and LLMs.

1. *Step 1 Warning signs:* The best system was the **high** precision scoring system using LLaMA 3 with a temperature of 0.1, achieving a weighted F1-score of 79.12%.
2. *Step 2 Internal coping strategies*: The best system was the **low-to-moderate** precision scoring system using O3-Mini with medium reasoning effort, achieving a weighted F1-score of 81.13%.
3. *Step 6 Making environment safe:* The best system was the **low-to-moderate** precision scoring system using O3-Mini with medium reasoning effort, achieving a weighted F1-score of 76.56%.
4. *Step 7 Reasons for living*: The best system was the **high** precision scoring system using O3-Mini with medium reasoning effort, achieving a weighted F1-score of 91.99%.

The confusion matrix in **Figure 5** illustrates how well the top-performing models’ predicted scores align with the raters’ scores across different categories. For *Step 1 warning signs* responses with the **high** precision scoring system, among those scored as 0 by professionals, 54% were correctly predicted as 0 by the LLM model, while 31% were misclassified as 1, and 14% were misclassified as 2. Among responses scored as 1, 88% were correctly predicted as 1, while 12% were misclassified as 2. Among responses scored as 2, 76% were correctly predicted as 2, while 1% were misclassified as 0, and 23% were misclassified as 1.

**Figure 5.**
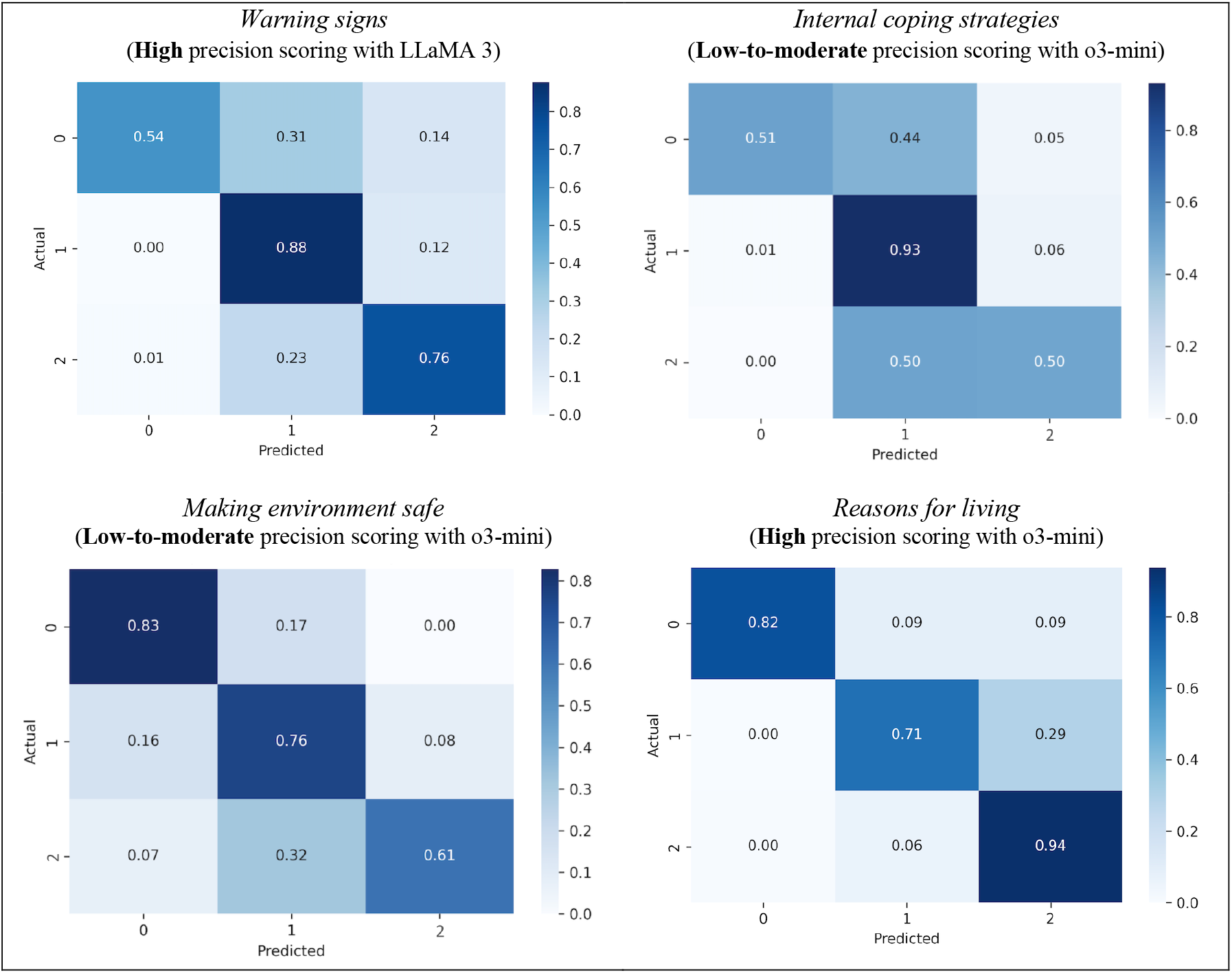
Confusion matrix of the best LLM models

For *Step 2 internal coping strategies* responses with the **low-to-moderate** precision scoring system, among those scored as 0 by professionals, 51% were correctly predicted as 0 by the LLM model, while 44% were misclassified as 1, and 5% were misclassified as 2. Among responses scored as 1, 93% were correctly predicted as 1, while 1% were misclassified as 0, and 6% were misclassified as 2. Among the responses rated as 2, half were correctly predicted as 2, while the other half were misclassified as 1.

For *Step 6 making environment safe* responses with the **low-to-moderate** precision scoring system, among those scored as 0 by professionals, 83% were correctly predicted as 0 by the LLM model, while 17% were misclassified as 1. Among responses scored as 1, 76% were correctly predicted as 1, while 16% were misclassified as 0, and 8% were misclassified as 2. Among responses scored as 2, 61% were correctly predicted as 2 and 7% were misclassified as 0, and 32% were misclassified as 1.

For *Step 7 reasons for living* responses with the **high** precision scoring system, among those scored as 0 by professionals, 82% were correctly predicted as 0 by the LLM model, while 9% were misclassified as 1, and another 9% were misclassified as 2. Among responses scored as 1 by professionals, 71% were correctly predicted as 1, while 29% were misclassified as 2. Among responses scored as 2, 94% were correctly predicted as 2 and 6% were

The results across different steps indicate variations in precision and recall for each score category (**Figure 6**). In Step 1 *warning signs*, Score 0 demonstrated moderate precision (0.79) and low recall (0.54); Score 1 had low precision (0.45) yet high recall (0.88); and Score 2 had high precision (0.95) with moderate recall (0.76). In Step 2 *internal coping strategies*, Score 0 achieved high precision (0.81) but had low recall (0.51); Score 1 showed both high precision (0.85) and recall (0.93); and Score 2 showed both low precision (0.67) and recall (0.50). In Step 6 *making environment safe*, Score 0 had a moderate precision (0.70) and high recall (0.83); Score 1 had high precision (0.85) and moderate recall (0.76); and Score 2 had low in both precision (0.54) and recall (0.61). In Step 7 *reason for living*, Score 0 had both high precision (0.95) and recall (0.82); Score 1 had low precision (0.44) but moderate recall (0.71); Score 2 had both high precision (0.97) and recall (0.94).

**Figure 6.**
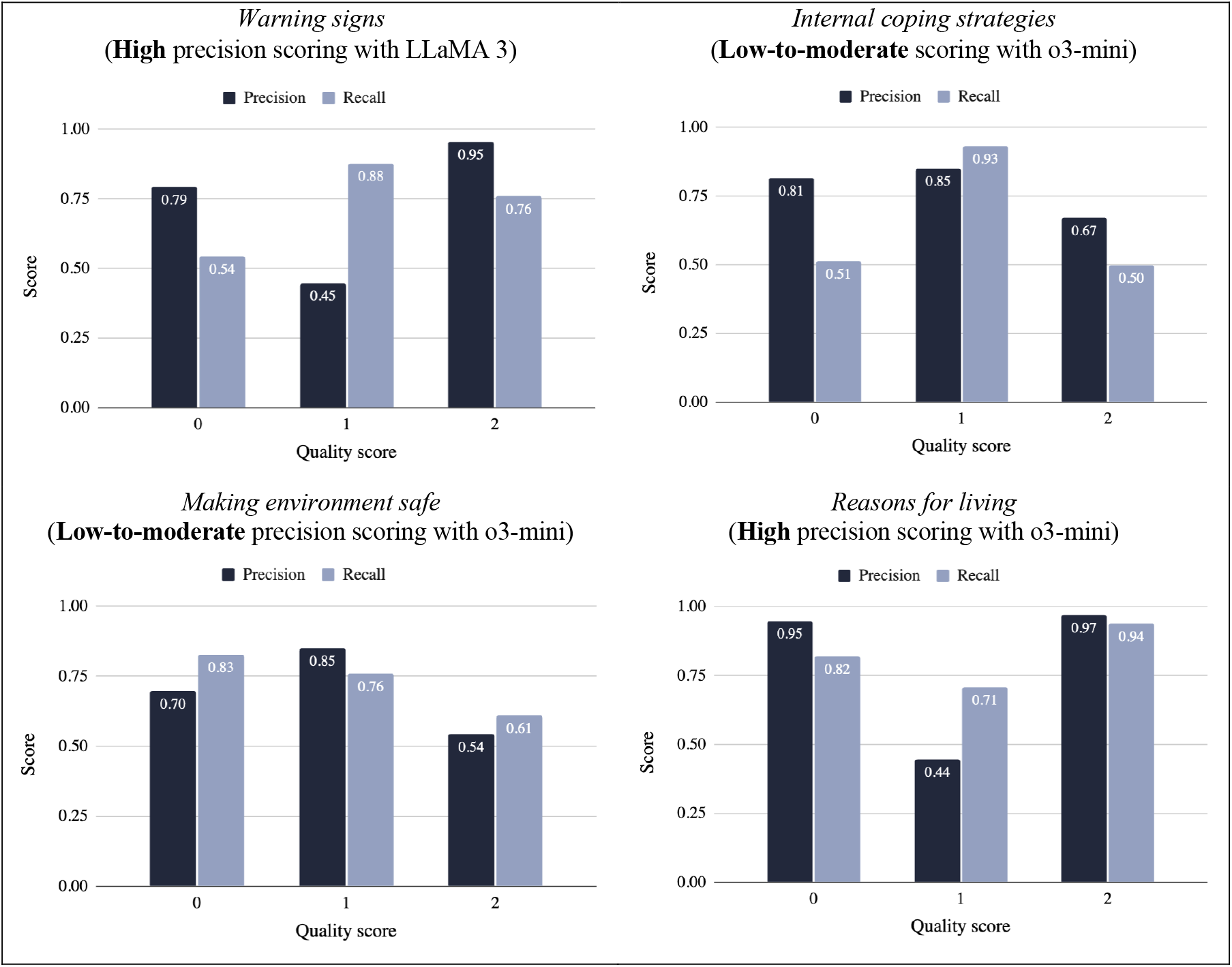
Precision and recall of the best LLM models

## Discussion

We assessed the performance of three LLMs for scoring safety plans. Overall, we observed that SPIFR accuracy improved when using the three-point scoring systems compared to the original four-point scoring system. We also learned that no one LLM provided the most optimal performance across steps and scoring systems. The current findings offer significant clinical implications, methodological advancements, and areas for future research.

From a clinical practice perspective, while existing LLM applications in suicide prevention focus on screening, diagnosis, or delivering eHealth services to patients^14, 15^, ours is one of the first to our knowledge that demonstrates the utility of LLM in assessing intervention quality. A deeper understanding of different LLM-base scoring systems and their clinical implications is essential for optimizing the provision of reliable, accurate feedback to clinicians. For example, although the **original** SPISA scoring method provided finely graded feedback, clinicians may struggle to distinguish subtle differences among scores. In contrast, the 3-point **high** scoring and **low-to-moderate** scoring systems may offer more reliable and distinguishable feedback, making it easier for clinicians to interpret score variations. A **high** scoring approach may be more appropriate when the fidelity tool aims to differentiate somewhat personalized responses from general ones, whereas **low-to-moderate** scoring is more effective in distinguishing highly personalized responses from somewhat personalized responses. Beyond predictive accuracy, future research should explore the relationship between different scoring approaches, different implementation strategies for providing feedback to clinicians and clinical outcomes (e.g., reducing patient suicide risk). The ultimate goal of this line of research is to establish that the automatic scoring system is optimally designed to enhance SPI effectiveness. Incorporating qualitative evaluations—such as clinicians’ agreement with LLMs’ reasoning—can further improve the interpretability and acceptability of AI-generated feedback. Furthermore, embedding this tool within the electronic health records systems will enable direct integration with patient-recorded safety plans in the patient medical record portal, ensuring ongoing engagement by providing timely and actionable feedback.

From a methodological perspective, this study is a first step in a scientific inquiry of important engineering questions to consider when designing an automatic scoring tool, such as the SPIFR, using medical record data. Beyond selection of which LLMs to assess, how to set the scoring systems, and which are most consistent in their ratings, other potential considerations include: 1) selecting one LLM model for all 7 steps of SPI or select the best model for each step, 2) creating an LLM ensemble-based voting approach to assigning a single score for each step, and 3) introducing few-shot learning and emerging methodologies to optimize LLM performance.

This study has several limitations. Generalizability of the study findings is limited to outpatient behavioral health settings and may not apply to inpatient hospital settings that are likely to include patients at higher risk for suicide. Another limitation involves the use of a selected sample, which consisted of safety plans that were not handwritten and were closely aligned with the Stanley-Brown Safety Plan form. Lastly, we compared three different LLMs; however, the significance of their performance variations has not been assessed. To validate these findings, additional statistical tests, such as McNemar’s Test, are recommended to determine whether the observed differences are statistically significant.

## Related works

This study is one of few, but critical emerging works in automated methods for scoring, characterizing, and assessing the efficacy of completed safety plans. Boggs et al^16^ developed a natural language processing (NLP) and rules-based system based on the ConText algorithm for identifying documented *professional contacts, lethal means counseling for firearms*, and *lethal means counseling for medication access/storage* from safety plans. Our study builds upon this work as the first study in applying LLMs to automatically score the quality of safety plans.

## Conclusion

From this pilot project, we conclude that LLMs have the potential to support an automatic SPIFR system and have identified clear paths toward improving LLM scoring performance and SPIFR methodological development.

## Data Availability

All data produced in the present study are only available upon reasonable request to the authors.

## Acknowledgement

This study was supported by the National Institute of Mental Health P50 MH127511 (Drs. Brown and Oquendo, MPIs) and R01 MH112139 (Dr. Stanley, PI).

